# Social networking service, patient-generated health data, and population health informatics: patterns and implications for using digital technologies to support mental health

**DOI:** 10.1101/2021.06.11.21258777

**Authors:** Jiancheng Ye, Zidan Wang, Jiarui Hai

## Abstract

**Objective:** To describe and compare characteristics of the population with and without mental health issues (depression or anxiety disorder), including physical health, sleep, and alcohol use. We also examined the patterns of social networking service use, patient-generated health data on the digital platforms, and health information sharing attitudes and activities.

**Methods:** We drew data from the National Cancer Institute’s 2019 Health Information National Trends Survey (HINTS). Participants were divided into two groups by mental health status. Then, we described and compared the characteristics of social determinants of health, health status, sleeping and drinking behaviors, and patterns of social networking service use and health information data sharing between the two groups. Multivariable logistic regression models were applied to assess the predictors of mental health. All analyses were weighted to provide nationally representative estimates.

**Results:** Participants with mental health issues are significantly more likely to be younger, White, female, have a lower income, have a history of chronic diseases, less capable of taking care of their own health; regarding behavioral health, they sleep less than six hours on average, have worse sleep quality, consume more alcohol; meanwhile, they are more likely to visit and share health information on social networking sites, write online diary blogs, participate online forum or support groups, watch health-related videos.

**Discussion and Conclusion:** This study illustrates that individuals with mental health issues have inequitable social determinants of health, poor physical health, and behavioral health. However, they are more likely to use social network platforms and services, share their health information, and have active engagements with patient-generated health data (PGHD). Leveraging these digital technologies and services could be beneficial to develop tailored and effective strategies for self-monitoring and self-management, thus supporting mental health.

## INTRODUCTION

Mental health issues, such as depression and anxiety disorder, are severe psychiatric diseases with high prevalence and elevated risks for recurrence and chronicity.[1] Globally, more than 260 million people of all ages have suffered from mental illnesses, which is a leading cause of disability worldwide and is a major contributor to the overall global burden of disease.[2] Studies have demonstrated that mental health issue is a strong indicator of poor general health, unhealthy alcohol use, and sleep problems.[3, 4] Poor sleep quality has been linked to an increased endorsement of drinking motives, especially for young adults.[5] It is critical for these patients to receive health care and social services capable of providing treatment and social support.

However, long-standing problems have hampered the efforts to improve mental health care delivery, quality of care, and social support. For example, mental health conditions are assessed exclusively on patients’ self-reporting, which could be burdensome to collect and subjective for clinical decision support. Currently, mental health services are mainly provided at times chosen by the practitioner rather than at the person’s time of greatest need.[6] The ideal way is to conduct regular assessments, which is useful to capture temporal dynamics of symptoms and crucial for both diagnosis and treatment planning. However, this could introduce burnout to both health care providers and patients.[7]

The emerging health technologies and digital services provide effective ways of collecting human behavior information, gathering patient-generated health data (PGHD), and sharing health-related information outside of clinical settings in a systematic way, and thus making interventions timely. Coupled with population health informatics tools, these technologies can track people’s digital exhaust, which includes PGHD and social network platform use. The rich real-time data enable researchers to gain insights into aspects of behavior that are well-established building blocks of mental health and illness, such as mood, social communication, sleep, alcohol use, and physical activity.

This study aims to describe and compare characteristics of the population with and without mental health issues (depression or anxiety disorder), including physical health, sleep, and alcohol use. We will also examine the patterns of social networking service use, patient-generated health data on the digital platforms, and health information sharing attitudes and activities. Figure 1 demonstrates the conceptual model - PBMI (**P**hysical and **B**ehavioral health, **M**ental health, and **I**nformatics) framework for this study. The results could provide a comprehensive understanding of the relationship among health, behavior, and informatics, which could be useful to develop tailored and effective strategies to support mental health management.

**Figure 1.**
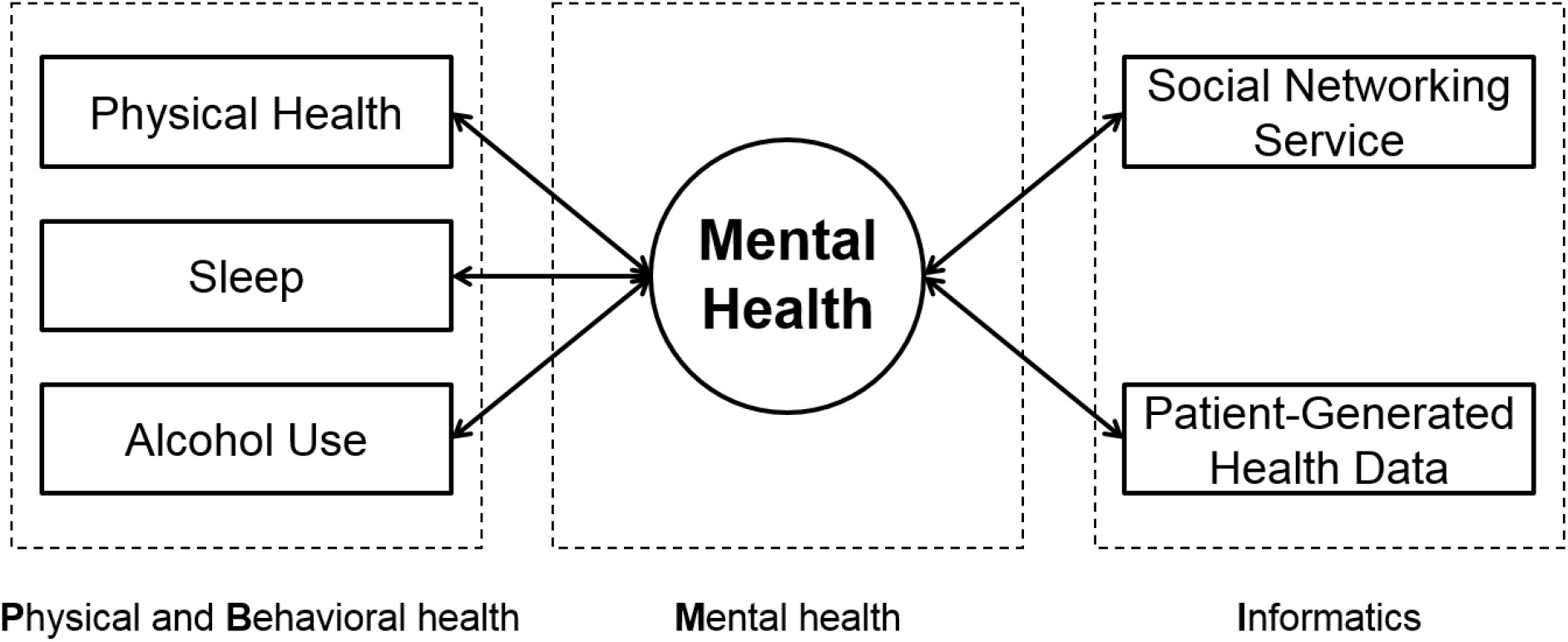
PBMI (**P**hysical and **B**ehavioral health, **M**ental health, and **I**nformatics) Framework

## METHODS

### Study design

Data for this study were drawn from the National Cancer Institute’s 2019 Health Information National Trends Survey (HINTS). HINTS is a nationally representative survey administered every year by the National Cancer Institute, which provides a comprehensive assessment of the American public’s current access to and use of health information.[8] The HINTS target population is civilian, non-institutionalized adults aged 18 or older living in the United States. In this study, we investigated the relationship among mental health, physical health, behavioral health, and social networking service.

### Study participants

Data used in this study were from the third round of data collection for HINTS 5 (Cycle 3), which was conducted from January 22 to April 30, 2019. Cycle 3 received 5,590 questionnaires, of which 5,438 were determined to be eligible after excluding blank, incomplete, and duplicate returned surveys.

In this study, the primary outcome is mental health, which is determined by the participant’s status of depression or anxiety disorder based on the results of the question “Has a doctor or other health professional ever told you that you had depression or anxiety disorder (yes/no)?” Among the 5,438 eligible respondents, 1,139 reported “yes,” 4,168 reported “no,” and 131 (2.41%) were missing and omitted in our analyses.

### Measures

#### Social determinants of health

The sample was divided into two groups by mental health status. Participants with depression or anxiety disorders were referred to as the group with mental health issues, and the others were referred to as the group with no mental health issues. We used participants’ self-reported information on age, sex, race, ethnicity, level of education, income, and usual source of care as our sociodemographic variables. We transformed the continuous variable age into a categorical variable by classifying age into four groups: (1) 18-34, (2) 35-49, (3) 50-64, and (4) 65+. Education level was re-categorized as less than college (including the post-high-school training), some college, college graduate and postgraduate degree; income level was re-categorized as ≤ 20,000, 20,000∼35,000, 35,000∼50,000, 50,000∼75,000, and > 75000 USD. We examined the participants’ history of chronic conditions on the four questions (all with yes or no responses): Has a doctor or other health professionals ever told you that you had (1) diabetes, (2) high blood pressure, (3) a heart condition, and (4) chronic lung disease?

#### Health-related information

To assess the participants’ general health, we considered their answers to questions related to physical health and mental status, including the ability to take care of health, emotion control by changing the way of thinking, and future consideration. We also included Patient Health Questionnaire-4 (PHQ-4), which was a derived composite from participants’ responses to questions on lack of interest in doing things, presence of depressed feelings, nervousness and anxiousness, and uncontrolled worry.[9]

#### PGHD and Social networking service

We examined participants’ use of the internet for health-related reasons from the five questions (all with yes or no responses): In the past 12 months, have you used the internet to (1) visit a social networking site, such as Facebook or LinkedIn, (2) share health information on social networking sites, such as Facebook or Twitter, (3) write in an online diary or blog, (4) participate in an online forum or support group for people with similar health or medical issue, and (5) watch a health-related video on YouTube?

We also inspected the first source of health information of the participants using the responses to the question “The most recent time you looked for information about health or medical topics, where did you go first?”, where a respondent can select one from twelve options. We further grouped the options into five main categories: internet, health professionals, family and friends, print materials, and others. In addition, we investigated participants’ attitudes to sharing health information, such as avoidance of doctor visits, talking about health with family and friends.

#### Alcohol consumption and sleep

We examined the participants’ alcohol consumption using the two questions: the number of days with at least one alcoholic drink per week and the average drinks per day. We assessed the participants’ sleep hours and quality using two questions: the average number of hours of sleep per night and the self-rated overall sleep quality. We transformed the continuous variable of average sleep per night into a categorical variable by classifying sleep hours to (1) 0-6 hours, (2) 7-8 hours, and (3) 9+ hours.

### Statistical Analysis

We used the survey package in R programming language to account for the complex sampling design used in HINTS and incorporated the Taylor series (linear approximation) [10] to generate accurate variance estimation. All analyses used weighted data to provide nationally representative estimates. The pairwise deletion was used to deal with missing data to preserve more information.

We compared mental health issues based on sample characteristics using the R version 4.0.5 (R Foundation, Vienna, Austria). To assess the characteristics of sociodemographic, general health, chronic diseases, social networking use, alcohol consumption, and sleeping variables, we generated weighted 2-way crosstab tables, which were tested with a Pearson Chi-square test of association.

The univariate logistic regression was built to examine the association between each predictor and mental health. Then we used multivariate logistic regression analyses using survey-weighted generalized linear modeling function in R. Odds ratio (OR) and 95% confidence interval (CI) for both models were presented. All reported P-values were two-tailed and the cutoff of P<0.05 was used to determine statistical significance for all analyses.

## RESULTS

### Population characteristics

Table 1 reports the characteristics of participants’ sociodemographic and clinical characteristics. Respondents with mental health issues were significantly more likely to be younger (P=0.004), White (P=0.005), female (P<0.001), have a lower income (<0.001), and have a usual source of care (P<0.001). They were also more likely to have a history of diabetes (P=0.001) and lung disease (P<0.001). There were no significant differences between the two groups regarding the characteristics of the history of hypertension (P=0.10), heart condition (P=0.40), and cancer (P=0.13).

**Table 1.** Unweighted and weighted prevalence estimates for sample sociodemographic, Health Information National Trends Survey (HINTS) 5 Cycle 3

| Characteristics | Overall<br>Unweighted %<br>(N=5,438) | Overall<br>Weighted %<br>(N=252,070,495) | Having mental health<br>issue, Weighted %<br>(N=57,953,433) | No mental health<br>issue, Weighted %<br>(N=189,456,090) | P-Value |
| --- | --- | --- | --- | --- | --- |
| **Age, %** |  |  |  |  | 0.004 |
| 18–34 | 13.0 | 24.3 | 28.1 | 23.1 |  |
| 35–49 | 18.3 | 24.5 | 26.3 | 24.2 |  |
| 50–64 | 31.6 | 31.1 | 32.8 | 30.7 |  |
| 65+ | 37.1 | 20.2 | 12.8 | 22.0 |  |
| **Male, %** | 42.1 | 48.8 | 37.9 | 52.5 | <0.001 |
| **Race\*, %** |  |  |  |  | 0.005 |
| White | 73.9 | 77.0 | 82.9 | 75.0 |  |
| Black | 16.5 | 13.0 | 9.9 | 13.9 |  |
| Asian | 5.0 | 5.8 | 3.4 | 6.7 |  |
| Others | 4.7 | 4.2 | 3.9 | 4.4 |  |
| **Hispanic ethnicity, %** | 14.9 | 16.7 | 13.9 | 17.6 | 0.07 |
| **Education, %** |  |  |  |  | 0.41 |
| High school diploma or less | 54.4 | 70.5 | 72.5 | 69.7 |  |
| College degree | 26.5 | 17.3 | 16.1 | 17.8 |  |
| Postgraduate | 19.1 | 12.2 | 11.4 | 12.5 |  |
| **Income, %** |  |  |  |  | <0.001 |
| Less than \$20,000 | 18.8 | 18.5 | 26.2 | 15.6 | |
| \$20,000–\$34,999 | 12.8 | 11.0 | 11.6 | 10.7 | |
| \$35,000–\$49,999 | 13.1 | 13.5 | 14.2 | 13.4 | |
| \$50,000–\$74,999 | 17.7 | 17.4 | 16.8 | 17.7 | |
| \$75,000 or more | 37.6 | 39.6 | 31.3 | 42.6 | |
| **Insurance, %** |  |  |  |  | 0.18 |
| Public | 44.1 | 35.2 | 38.9 | 33.5 |  |
| Private | 42.3 | 48.9 | 44.8 | 50.8 |  |
| Uninsured | 4.8 | 7.7 | 7.2 | 7.8 |  |
| Others** | 8.8 | 8.2 | 9.2 | 8.0 |  |
| **Has a usual source of care, %** | 69.8 | 64.5 | 73.7 | 61.7 | <0.001 |
| **Has cancer history, %** | 16.1 | 9.5 | 7.9 | 9.7 | 0.13 |
| **History of lung disease, %** | 11.8 | 11.2 | 21.0 | 8.2 | <0.001 |
| **History of heart condition, %** | 16.1 | 8.1 | 9.2 | 7.8 | 0.40 |
| **History of diabetes, %** | 21.7 | 17.0 | 22.4 | 15.2 | 0.001 |
| **History of hypertension, %** | 45.0 | 36.0 | 39.1 | 34.9 | 0.10 |

**Note:** \*Race: Asian Indian, Chinese, Filipino, Japanese, Korean, Vietnamese, and other Asian were collapsed into the “Asian” category; race categories other than White, Black, and Asian were reclassified as “Others”.
\*\* Insurance: “Others” includes coverage under the spouse, coverage under parents, low-income beneficiary.

### Health information and social networking service

Table 2 shows the characteristics of health information source, sharing, and social networking service use. Mental health patients are more likely to have worse general health status (P<0.001), less confidence in taking care of their own health (P<0.001), and have a larger PHQ4 score (P<0.001). Individuals with mental health issues also tend to be less likely to control emotions by changing the way they are thinking about the situation (P<0.001) or try to influence things in the future with day-to-day behavior (P<0.001). In addition, Table 2 shows that those with mental health issues are significantly more likely to visit social networking sites (P=0.04), share health information on social networking sites (P=0.001), write in an online diary blog (P=0.007), participate in an online forum or support group (P<0.001), watch health-related videos (P=0.009). There are no significant differences in the first source of health information (P=0.225), using wellness apps (P=0.33), avoidance of doctor visits (P=0.151), and talking about health with family or friends (P=0.076) between the two groups.

**Table 2.** Prevalence estimates for characteristics of health information and social networking service

| Characteristics | Overall<br>Unweighted %<br>(N=5,438) | Overall<br>Weighted %<br>(N=252,070,495) | Having mental<br>health issue<br>Weighted %<br>(N=57,953,433) | No mental<br>health issue<br>Weighted %<br>(N=189,456,090) | P-Value |
| --- | --- | --- | --- | --- | --- |
| **Source of health information, %** |  |  |  |  | 0.23 |
| Internet | 42.9 | 46.1 | 50.2 | 45.2 |  |
| Health professionals | 48.9 | 44.6 | 39.9 | 45.9 |  |
| Family or friends | 4.1 | 5.2 | 4.6 | 5.3 |  |
| Print materials | 2.3 | 2.2 | 2.4 | 2.1 |  |
| Others | 1.8 | 2.0 | 2.9 | 1.5 |  |
| **Use health apps, %** |  |  |  |  | 0.33 |
| Yes | 52.4 | 54.8 | 57.7 | 54.2 |  |
| No | 42.2 | 39.6 | 35.9 | 40.6 |  |
| Don't Know | 5.4 | 5.5 | 6.4 | 5.2 |  |
| **Good health status, %** | 47.9 | 49.4 | 33.7 | 54.2 | < 0.001 |
| **Have ability to take care of health, %** | 72.2 | 71.5 | 56.0 | 76.3 | <0.001 |
| **Avoid visiting doctor, %** | 25.0 | 30.6 | 33.9 | 29.6 | 0.15 |
| **Talks about health with family or friends, %** | 81.0 | 78.5 | 81.9 | 77.4 | 0.08 |
| **PHQ 4,%** |  |  |  |  | <0.001 |
| 0 | 50.3 | 45.6 | 13.6 | 54.9 |  |
| 1 | 12.3 | 13.2 | 9.3 | 14.4 |  |
| 2 | 9.9 | 9.6 | 11.5 | 9.1 |  |
| 3+ | 27.6 | 31.6 | 65.5 | 21.6 |  |
| **Can control emotions, %** | 85.1 | 84.5 | 77.6 | 86.7 | <0.001 |
| **Consider future, %** | 84.5 | 84.7 | 79.2 | 86.6 | <0.001 |
| **Visit social networking sites, %** | 65.0 | 71.5 | 76.0 | 70.8 | 0.04 |
| **Share health information, %** | 11.9 | 14.6 | 19.7 | 13.1 | 0.001 |
| **Write online diary or blog, %** | 3.6 | 5.0 | 8.0 | 4.0 | 0.007 |
| **Participate online forum or health-related group, %** | 7.0 | 8.1 | 13.3 | 6.6 | <0.001 |
| **Watch health-related videos, %** | 32.8 | 37.3 | 42.5 | 35.9 | 0.009 |

### Alcohol consumption and sleep

Table 3 shows the characteristics of behavioral health, including sleep and alcohol use, between the two groups. Individuals with mental health issues are more likely to sleep less than 6 hours or more than 9 hours (P=0.01), have worse sleep quality (P<0.001), and consume more alcohol per day (P=0.03). There was no difference in the number of days having alcohol per week between the two groups.

**Table 3.**
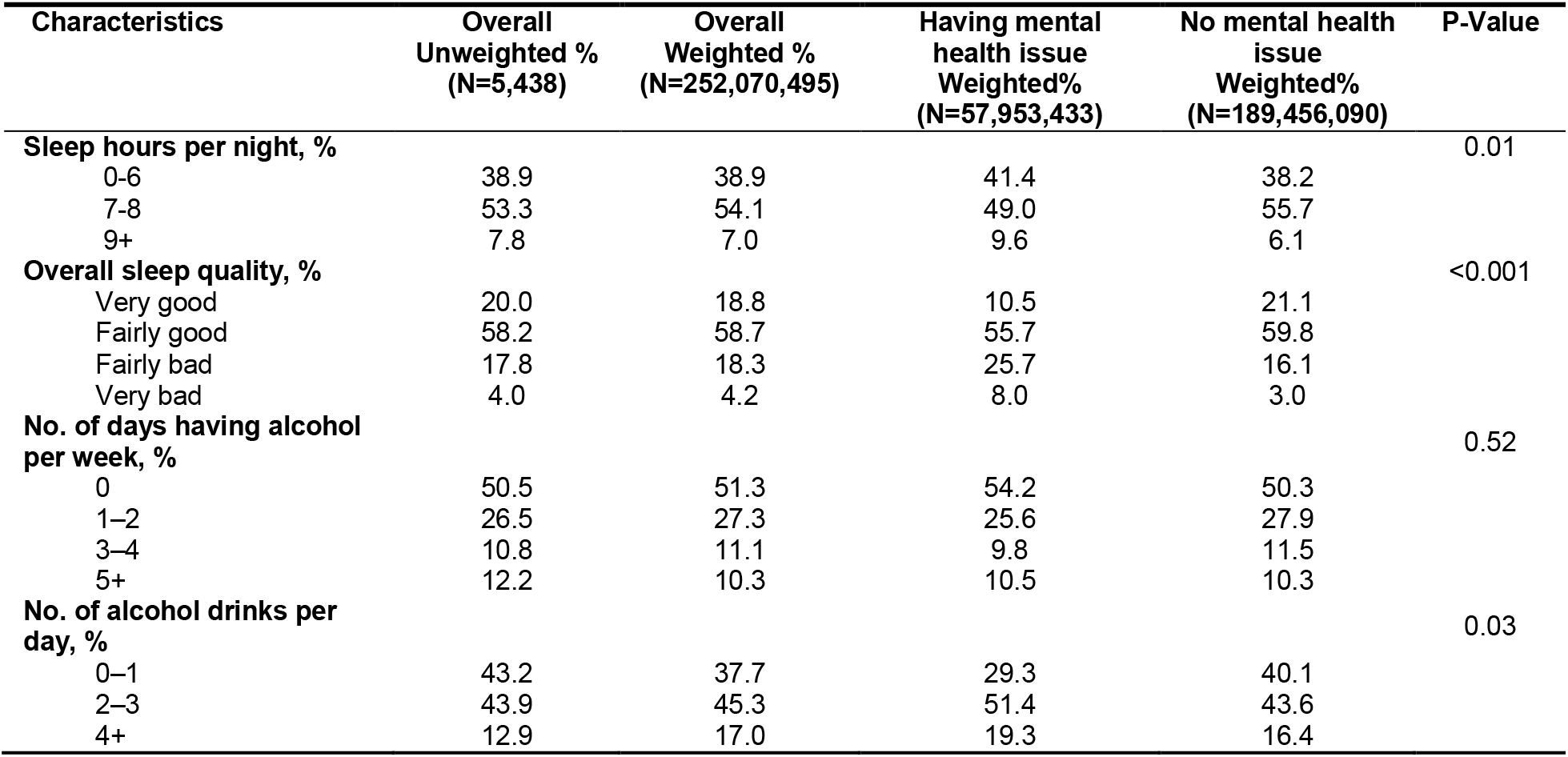
Prevalence estimates for characteristics of sleep and alcohol use

| Characteristics | Overall<br>Unweighted %<br>(N=5,438) | Overall<br>Weighted %<br>(N=252,070,495) | Having mental<br>health issue<br>Weighted%<br>(N=57,953,433) | No mental health<br>issue<br>Weighted%<br>(N=189,456,090) | P-Value |
| --- | --- | --- | --- | --- | --- |
| **Sleep hours per night, %** |  |  |  |  | 0.01 |
| 0-6 | 38.9 | 38.9 | 41.4 | 38.2 |  |
| 7-8 | 53.3 | 54.1 | 49.0 | 55.7 |  |
| 9+ | 7.8 | 7.0 | 9.6 | 6.1 |  |
| **Overall sleep quality, %** |  |  |  |  | <0.001 |
| Very good | 20.0 | 18.8 | 10.5 | 21.1 |  |
| Fairly good | 58.2 | 58.7 | 55.7 | 59.8 |  |
| Fairly bad | 17.8 | 18.3 | 25.7 | 16.1 |  |
| Very bad | 4.0 | 4.2 | 8.0 | 3.0 |  |
| **No. of days having alcohol**<br>**per week, %** |  |  |  |  | 0.52 |
| 0 | 50.5 | 51.3 | 54.2 | 50.3 |  |
| 1-2 | 26.5 | 27.3 | 25.6 | 27.9 |  |
| 3-4 | 10.8 | 11.1 | 9.8 | 11.5 |  |
| 5+ | 12.2 | 10.3 | 10.5 | 10.3 |  |
| **No. of alcohol drinks per**<br>**day, %** |  |  |  |  | 0.03 |
| 0-1 | 43.2 | 37.7 | 29.3 | 40.1 |  |
| 2-3 | 43.9 | 45.3 | 51.4 | 43.6 |  |
| 4+ | 12.9 | 17.0 | 19.3 | 16.4 |  |

Figure 2 illustrates the difference in sleep quality among individuals who sleep no more than 6, from 7 to 8, and more than 9 hours per night between the two groups, respectively. For individuals with mental health issues, 52% of the individuals who sleep no more than 6 hours per night have poor sleep quality. Among individuals without mental health issues, only 9.1% of those who sleep 7 to 8 hours per night have a poor sleep quality, which is significantly less than individuals with mental health issues who sleep the same hours.

**Figure 2.**
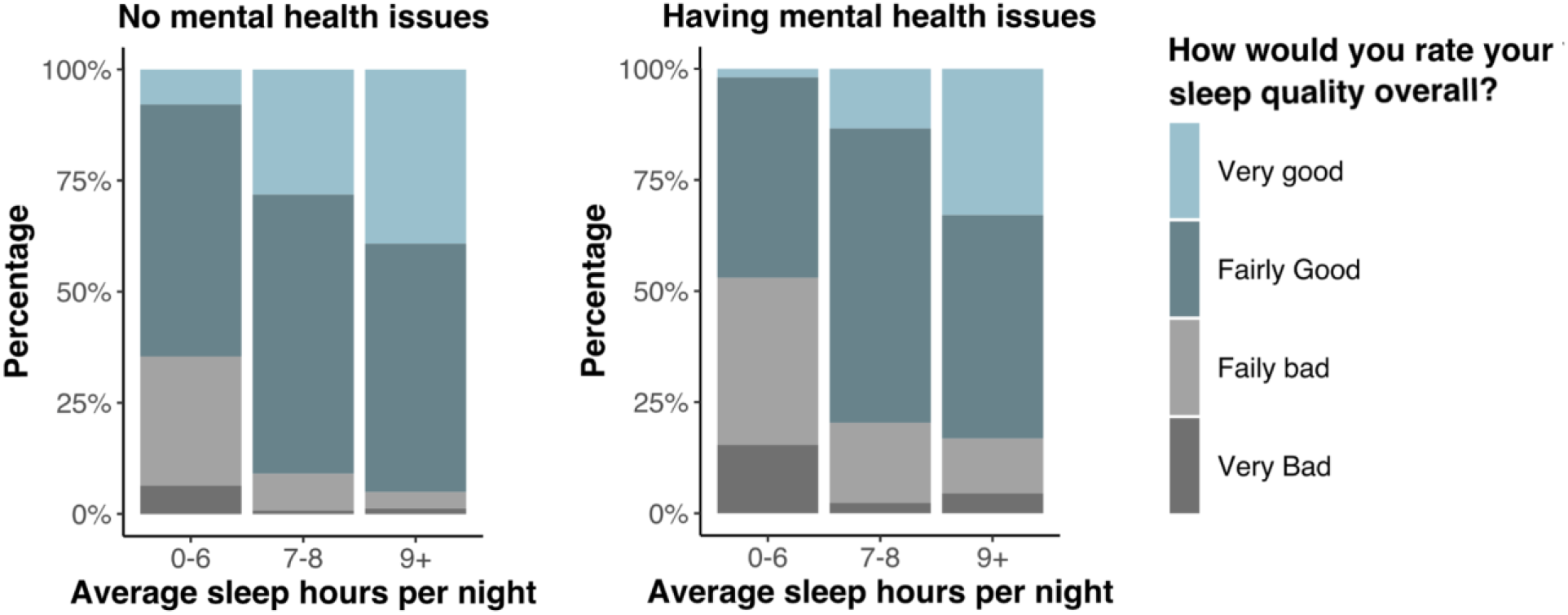
Sleep patterns between two mental health groups

We examined the pattern that whether the sleep quality is identical for people with and without mental health issues separately for the three sleep hour categories. Since the normality assumption is unjustified, we conducted the Mann-Whitney U test. For people who sleep 0-6 (P<0.001) and 7-8 hours (P<0.001), there is a significant difference in quality between the two groups. There is no significant difference in quality for people who sleep more than 9 hours (P=0.14) between the two groups. In general, individuals without mental health issues sleep 7-8 hours with higher quality, while mental health patients sleep less than 6 or more than 9 with poor quality.

Figure 3 illustrates the difference in the amount of alcohol consumed per week among groups stratified by sex and mental health status. 43% of the females without mental health issues consume one or more drinks per week, while 44.8% of the females with mental health issues consume the same number of drinks per week. We found no significant difference in drink amount between females (P=0.66) and males (P=0.23) regardless of mental health status. However, for individuals without mental health, males drink significantly more than females (P<0.001).

**Figure 3.**
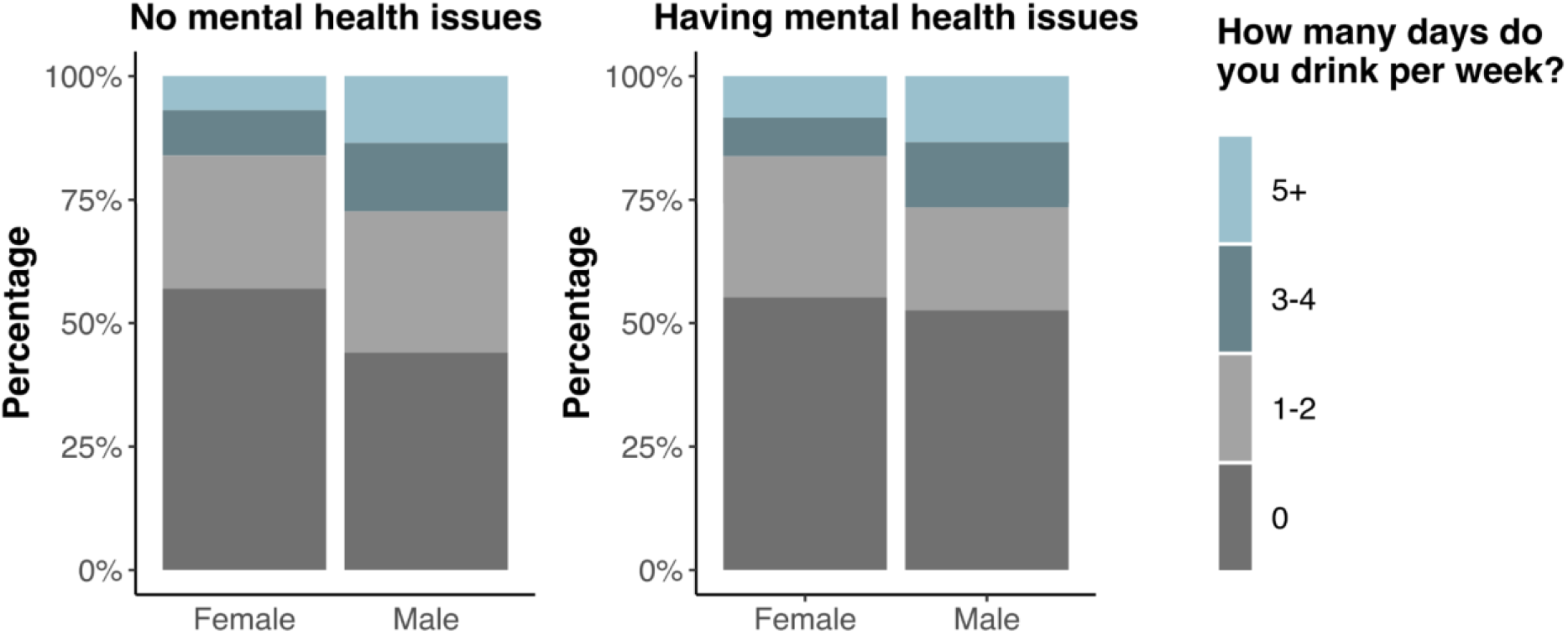
Alcohol use patterns by sex and mental health

### Social determinants of health and mental health

Table 4 shows the results of logistic regression analyses. In the unadjusted logistic regression model, most covariates are associated with mental health. In the adjusted model, those aged 65+ have a reduced likelihood (OR 0.20; 95% CI: 0.11-0.35) of having mental health issues, compared to those aged 18-34. Males are less likely (OR 0.52; 95% CI: 0.40-0.68) to have mental health issues. The Black population has a reduced likelihood (OR 0.41; 95% CI: 0.27-0.63) of having mental health issues compared to the White population.

**Table 4.** Crude and adjusted odds from logistic regression analyses of associations between social determinants of health and mental health

| Predictor | Unadjusted |  | Adjusted |  |
| --- | --- | --- | --- | --- |
|  | OR (95% CI) | P-value | OR (95% CI) | P-value |
| **Age** |  |  |  |  |
| 18-34 | Reference |  | Reference |  |
| 35-49 | 0.89 (0.61, 1.31) | 0.56 | 0.99 (0.64, 1.54) | 0.98 |
| 50-64 | 0.88 (0.60, 1.29) | 0.51 | 0.60 (0.38, 0.96) | 0.04 |
| 65 and older | 0.48 (0.32, 0.71) | <0.001 | 0.20 (0.11, 0.35) | <0.001 |
| **Sex** |  |  |  |  |
| Male | 0.55 (0.43, 0.70) | <0.001 | 0.52 (0.40, 0.68) | <0.001 |
| **Race** |  |  |  |  |
| White | Reference |  | Reference |  |
| Asian | 0.46 (0.25, 0.86) | 0.02 | 0.51 (0.25, 1.03) | 0.06 |
| Black | 0.64 (0.46, 0.90) | 0.01 | 0.41 (0.27, 0.63) | <0.001 |
| Others | 0.79 (0.48, 1.32) | 0.37 | 0.52 (0.25, 1.08) | 0.08 |
| **Ethnicity** |  |  |  |  |
| Hispanic | 0.76 (0.56, 1.02) | 0.07 | 0.73 (0.48, 1.11) | 0.14 |
| **Education** |  |  |  |  |
| High school diploma or less | Reference |  | Reference |  |
| College degree | 0.87 (0.68, 1.12) | 0.28 | 0.91 (0.64, 1.31) | 0.63 |
| Postgraduate | 0.87 (0.65, 1.17) | 0.37 | 0.92 (0.59, 1.42) | 0.69 |
| **Income** |  |  |  |  |
| Less than \$20,000 | 2.29 (1.65, 3.17) | <0.001 | 2.39 (1.48, 3.84) | <0.001 |
| \$20,000–\$34,999 | 1.48 (1.00, 2.19) | 0.05 | 1.59 (1.00, 2.55) | 0.05 |
| \$35,000–\$49,999 | 1.44 (0.97, 2.14) | 0.07 | 1.34 (0.83, 2.15) | 0.23 |
| \$50,000–\$74,999 | 1.29 (0.89, 1.86) | 0.18 | 1.32 (0.88, 1.98) | 0.19 |
| \$75,000 or more | Reference | | Reference | |
| **Insurance** |  |  |  |  |
| Public | Reference |  | Reference |  |
| Private | 0.76 (0.59, 0.98) | 0.03 | 0.71 (0.47, 1.07) | 0.11 |
| Uninsured | 0.79 (0.48, 1.31) | 0.37 | 0.56 (0.29, 1.09) | 0.09 |
| Others | 0.99 (0.65, 1.51) | 0.98 | 0.94 (0.59, 1.51) | 0.81 |
| **Has a usual source of care** | 1.74 (1.36, 2.24) | <0.001 | 1.72 (1.24, 2.39) | 0.001 |
| **Has cancer history** | 0.81 (0.61, 1.07) | 0.14 | 0.88 (0.61, 1.28) | 0.51 |
| **History of lung disease** | 2.99 (2.21, 4.04) | <0.001 | 2.17 (1.54, 3.05) | <0.001 |
| **History of heart condition** | 1.19 (0.79, 1.80) | 0.40 | 0.94 (0.60, 1.47) | 0.79 |
| **History of diabetes** | 1.61 (1.23, 2.10) | <0.001 | 1.42 (1.03, 1.95) | 0.03 |
| **History of hypertension** | 1.20 (0.97, 1.48) | 0.10 | 1.40 (1.07, 1.84) | 0.01 |

Individuals who have an annual family income less than $20,000 are more likely (OR 2.39; 95% CI: 1.48-3.84) to have mental health issues than those whose income is more than $75,000. Those having a usual source of care are more likely (OR 1.72; 95% CI: 1.24-2.39) to have mental health issues. As expected, those having a history of lung disease (OR 2.17; 95% CI 1.54-3.05), diabetes (OR 1.42; 95% CI 1.03-1.95), and hypertension (OR 1.40; 95% CI 1.07-1.84) are more likely to have mental health issues. The results also indicate that ethnicity, education, insurance type, cancer history, and history of heart condition have no association with mental health status.

## DISCUSSION

This study aims to describe and compare characteristics of the population with and without mental health issues (depression or anxiety disorder), including physical health, sleep, and alcohol use. We will also examine the patterns of social networking service use, patient-generated health data on the digital platforms, and health information sharing attitudes and activities. We found that participants who were younger, White, female, had a lower income, had a history of chronic disease, had a higher PHQ4 score, were more likely to have mental health problems, which was consistent with previous findings. [11]

Participants with mental illness were more likely to visit social networking sites, share health information on social networking sites, write in an online diary blog, participate in an online forum or support group, watch health-related videos. We also found that participants with mental illness slept less with worse sleep quality and consumed more alcohol per day.

Overall, social determinants of health like age, race, income, insurance status, and chronic diseases like lung disease, diabetes, and hypertension are associated with mental health. In recent years, there has been increasing acknowledgment of the important role mental health plays in achieving improved population health. Understanding how these fundamental factors (physical and behavioral health, mental health, and technologies) relate to one another may yield important insights for novel approaches in designing prevention programs and enhancing services for mental health support. Digital health technologies such as smartphone applications and social media provide opportunities to continuously collect objective information on behavior in the context of people’s real lives, generating a rich data set that can provide insight into the extent and timing of mental-health needs in individuals.[12]

### Social networking service

Individuals who have depression and anxiety are more likely to use social network platforms, especially younger people. They also tend to be less likely to control emotions by changing the way they are thinking about the situation or try to influence things in the future with day-to-day behavior. Social networking played an important role for this population to find ways to reduce loneliness or symptoms of mental health problems.

We also found that women had a higher level of vulnerability to poor mental health compared to men, which aligned with previous findings. [13] There is an ongoing debate about whether the use of mobile health technologies such as social media is detrimental to mental health.[14] Interestingly, those with depression or anxiety disorder were significantly more likely to visit social networking sites, write online diary blogs, participate in an online forum or support group, and watch health-related videos. Those social network platforms could potentially provide effective strategies to intervene in mental illness. We acknowledge that safe small to moderate use of social media is beneficial, but it could introduce harmful influences if people spend too much time in this digital and virtual world.[15] Further research is needed to understand the quantitative and dynamic patterns of social media use to measure the benefits versus harmful effects and inform evidence-based approaches to clinical interventions, practices, policy, education, and regulation.[16] If we take advantage of the social networking service and data-gathering functions of digital platforms in the right ways, we may achieve breakthroughs in the ability to take care of mental health and well-being.

### Mental health and patient-generated health data

This study found that individuals with depression or anxiety disorder were willing to share health information on social networking sites. They offer an opportunity to provide interventions that are timely, personalized, and scalable. Coupled with telehealth or remote management platforms, mental health services and support could be provided by the practitioners in a timely manner and at the person’s time of greatest need. The digital health platforms and PGHD are facilitating the development of a wave of timely interventions for mental health care and support.[6] These findings may be useful for stakeholders like mental health providers, researchers, public health practitioners, mobile health and social media companies to work jointly to design and provide “precision social networking service” with higher personalized and participatory levels, thus improving the population health.

### Mental health, alcohol use, and sleep

This study found that individuals without mental health issues sleep 7-8 hours with higher quality, while mental health patients sleep less than 6 or more than 9 hours with poor quality. Scientific guidelines for sleep suggest that 7 or more hours of sleep per night are appropriate for 18–60 years adults, 7–9 hours for 61–64 adults, and 7–8 hours for 65 years and older.[17, 18] Although the amount is important, other aspects of sleep also contribute to health and well-being. Good sleep quality is also essential. We also found that mental health patients were more likely to sleep too much, which was not recommended. Previous studies show that adolescents and young adults are prone to both mental health and sleep problems. [19] Sleep quality may be particularly important for young adults such as college students with poor mental health who, compared with peers, tend to lack protective social support networks.[20]

Among individuals who are already susceptible to alcohol use, inadequate sleep may further weaken their cognitive capacity to make safer drinking-related decisions or self-protective behaviors, irrespective of consumption levels. Further investigations are needed to examine how poor mental health relates to both alcohol consumption and consequences, as well as the extent to which alcohol consumption may mediate the relationship between mental health and consequences. Digital social platforms play a vital role in educating people on alternative coping skills or harm-reduction skills to use in drinking contexts.

Given the important role of different types of drinking motives in the connection between mental health and drinking outcomes, it is important to examine drinking motivations as mediators of this relationship.[21] Furthermore, event-level methods that simultaneously account for individuals’ sleep and alcohol use behaviors may be helpful for future longitudinal research.

### Limitation

The sample was confronted with missing data regarding health outcomes and covariates, which may not be missing completely at random. Those who did not respond to questions may be less active, and thus our estimates may be subject to bias. Because the survey was cross-sectional, we could not examine causality among variables. Meanwhile, given the limitations of the dataset, we did not have information about social network platforms’ usage frequency and duration. Despite these limitations, this study contributes to a better understanding of the effects and patterns among social networking services, PGHD, social determinants of health, and mental health.

## CONCLUSION

This study illustrates that social determinants of health are associated with mental health issues. Health disparities exist between women and men, and among different races, in mental health. Mental health issues result in less sleep with poor quality and unhealthy alcohol drinking behaviors. Individuals with mental health issues are more likely to use social networking platforms, share their health information, and have active engagements in patient-generated health data (PGHD). The results provide important insight into the interplay between three vital health-related domains—physical health, behavioral health (sleep and alcohol use), social networking service, and their patterns on the mental health patient population. Leveraging digital platforms and population informatics such as mobile health and social media along with PGHD could offer unique opportunities to develop effective self-monitoring and management strategies for supporting mental health patients.

## Data Availability

All data referred to in the manuscript are available on: https://www.cancer.gov/

## CONTRIBUTORSHIP STATEMENT

J.Y. conceived and designed the study, and contributed to the analyses. Z.W. contributed to the analyses. J.Y., Z.W., J.H. contributed to the interpretation of the results, drafting and revision on the manuscript. All the authors read and approved the final version of the manuscript.

